# Corticosteroid Responsiveness Following COPD Exacerbations with Respiratory Failure

**DOI:** 10.64898/2026.09.24.26363953

**Authors:** Jonathan K. Zakrajsek, Sung-Joon Min, Hyunmin Kim, Jihye Kim, Harold W. Bell, Fernando Diaz del Valle, Moumita Ghosh, Martin R. Zamora, Joyce S. Lee, Trevor C. Steinbach, Tzu L. Phang, Adit A. Ginde, Tyree H. Kiser, Anthony N. Gerber, R. William Vandivier

## Abstract

**Study Question:** COPD exacerbations with respiratory failure have an outsized impact on morbidity, mortality, and healthcare costs. Physicians treat these severe events with high doses of corticosteroids out of concern for intrinsic corticosteroid resistance. This study was designed to determine whether people who suffer from exacerbations with respiratory failure have intrinsic steroid resistance that could affect that decision.

**Patients and Methods:** A matched, case-control study was performed at University of Colorado. Subjects were enrolled into the Exacerbation group if they were hospitalized for an exacerbation requiring ventilation, or into the Stable COPD group if they had no inpatient exacerbation within 12 months. Stable COPD subjects were frequency-matched to Exacerbation subjects for age, smoking status and FEV_1_%. Corticosteroid sensitivity was assessed *ex vivo* and *in vivo* >45 days following the last exacerbation during a period of relative quiescence. The primary outcome was the group difference in methylprednisolone IC_50_ for LPS-stimulated IL-8 release by mononuclear cells.

**Measurements and Main Results:** The Exacerbation group (N=8) had more exacerbations in the prior year (6.0 vs. 0.5; p<0.01) and had higher baseline blood lymphocytes (p<0.01) *versus* Stable controls (N=8). *Ex vivo*, methylprednisolone equally suppressed IL-8, IL-6, TNFα and IL-1β release by mononuclear cells and whole blood across groups. *In vivo*, methylprednisolone decreased plasma mediators and mononuclear cells in both groups but had a small differential effect on mononuclear cell gene expression. **Answer to the Question**. Exacerbations with respiratory failure have persistent lymphocytosis, an altered gene response to corticosteroids, but no corticosteroid resistance to suppress inflammation.

## Introduction

Acute exacerbations of chronic obstructive pulmonary disease (exacerbations) are the most important complication of COPD, because they are common, herald a steep decline in quality of life, markedly increase the risk of death, and drive ~60% of the expenditure for COPD in the United States[1–6].

Consequences are particularly high for exacerbations with respiratory failure that require invasive or non-invasive ventilator support in the intensive care unit, because healthcare costs are increased[7] and hospital mortality rates can reach as high as ~25-30%[5, 8]. The long-term outlook is dismal following an exacerbation with respiratory failure, since 80% will be re-admitted, 63% will have another life-threatening exacerbation and ~60% will die within the next year[5, 8–12].

Standard therapies for exacerbations include bronchodilators, antibiotics and corticosteroids. Corticosteroids improve oxygenation, speed recovery, reduce treatment failure and have been recommended for exacerbations of moderate severity or greater[13–16]. Clinical trials suggest that most moderate to severe exacerbations that are not treated with invasive or non-invasive ventilation can be effectively treated with prednisone as low as 40mg per day for 5 days[13–15]. In contrast, many clinicians treat exacerbations with respiratory failure that require invasive or non-invasive ventilatory support with doses of corticosteroids that are up to 12 times higher, perhaps due to the perception that they have an underlying resistance to corticosteroids[7, 17, 18].

This study was designed to test the hypothesis that people with a recent exacerbation complicated by respiratory failure are resistant to corticosteroids and that this resistance is part of their underlying endotype. Exacerbation subjects were recruited to participate in the study during hospitalization if they were ≥ 40 years old, had an emergency department or intensive care unit diagnosis of an exacerbation and were treated with noninvasive or invasive ventilation within the first 24 hours of hospitalization. Exacerbation subjects returned for *ex vivo* and *in vivo* steroid sensitivity studies during a period of relative quiescence defined as > 45 days post hospitalization and > 30 days after their last treatment with antibiotics or corticosteroids. Results of steroid sensitivity studies were compared to identical studies performed on Stable COPD subjects who were frequency-matched to exacerbation subjects for age, FEV_1_ % predicted and current smoking status.

## Material and Methods

See online data supplement for detailed methods

### Study Participants

Exacerbation subjects were recruited during their index hospitalization and were invited to return for an outpatient visit for testing ≥45 days after hospital discharge. Matched, Stable COPD controls were recruited from the COPD and Lung Transplantation Clinics. Inclusion criteria for the exacerbation cohort were 1) emergency department or intensive care unit physician diagnosis of a COPD exacerbation, 2) age ≥ 40 years, 3) need for non-invasive or invasive ventilator support in the emergency department or intensive care unit during the first 24 hours of hospitalization. Inclusion criteria for the Stable COPD cohort were 1) physician diagnosis of COPD and 2) age ≥ 40 years. Stable COPD patients were also frequency-matched to exacerbation patients for age (± 10 year increments), current/former smoking status (former smoker = no smoking for ≥ 1 month) and lung function (FEV_1_% predicted by ± 10% increments). The study excluded people in the Stable COPD group if they had a moderate exacerbation treated as an outpatient or through the emergency department within 6 months, or a severe exacerbation treated as an inpatient within 12 months. Subjects in both cohorts were excluded for systemic steroid or antibiotic use ≤ 30 days prior to the return visit.

### Study Design

We conducted a matched, case-control study from October 2017 through June 2020 to test the hypothesis that people with a recent exacerbation complicated by respiratory failure are resistant to corticosteroids and that this resistance is a component of their underlying endotype. (ClinicalTrials.gov ID: NCT03680495). The Colorado Multiple Institutional Review Board approved the study (COMIRB 16-0256) and all subjects completed written, informed consent.

### Methods

The primary study outcome was steroid resistance measured by a difference in the *ex vivo* IC_50_ of methylprednisolone to suppress IL-8 release by peripheral blood mononuclear cells (mononuclear cells) stimulated with LPS in Exacerbation *versus* Stable COPD groups.

#### Ex Vivo Studies

Mononuclear cells or heparinized whole blood were treated with methylprednisolone at 0, 10^10^, 10^9^, 10^8^, 10^7^, 10^6^ and 10^5^ M (Sigma-Aldrich, Saint Louis, MO, 63103) for 1 hour and stimulated with *E. coli* O11B4 LPS (List Labs, Campbell, CA, 95008) at 10 ng/ml for 24h. Supernatants were assessed for IL-8, IL-6, TNFα and IL-1β (Meso Scale Diagnostics, Rockville, MD 20850) to determine the methylprednisolone IC_50_.

#### In Vivo Studies

*In vivo* studies of steroid resistance were performed by testing the ability of methylprednisolone to suppress pro-inflammatory cytokines (IL-8, TNFα, IL-6, IFN-γ, IL-5) and chemokines (MIP-3α, MCP-1, MCP-4, IP-10, Eotaxin), and to activate the anti-inflammatory mediator, IL-10 (Meso Scale Diagnostics, Rockville, MD 20850) in serum at 0 and 6 hours post intravenous injection with 60 mg of methylprednisolone (Sigma-Aldrich, Saint Louis, MO, 63103). Group differences on total mononuclear cells, monocyte and lymphocyte cell counts were assessed in mononuclear cells collected at 0, 2, 4 and 6 hours following intravenous methylprednisolone.

#### RNA Isolation and Sequencing

See online data supplement for detailed methods.

#### Clinical Measurements

See online data supplement for detailed methods.

### Analysis

Statistical analyses were performed using SAS for Windows, version 9.4 (SAS Institute, Cary, NC). Comparisons between study subjects in the Exacerbation and Stable COPD groups were made (using t-tests or Wilcoxon tests if the distributions were skewed) for continuous variables and chi-square tests (or Fisher’s exact tests if cell sizes were small) for categorical variables.

Comparisons involving longitudinal curves were carried out using linear models. All tests were 2-sided with a significance level of p < 0.05. All graphs show standard error.

## Results

Two hundred and fifty-five Exacerbation subjects were screened and 60 were eligible. Of these, 28 were enrolled and completed baseline testing during hospitalization (Fig. 1). Two enrolled subjects were removed after it was found that they had restrictive lung physiology and interstitial pneumonitis, respectively. Therefore, 26 Exacerbation subjects were included in the analysis. Eighteen Exacerbation subjects did not complete the follow up visit due to voluntary withdrawal (N=6), death or decision to seek hospice care (N=5), inability to re-establish contact (N=4) and continued steroid use (N=3). Eight Exacerbation subjects followed up for testing and completion of the study at 91.6 ± 43.5 days post hospitalization. One subject only completed questionnaires during follow up due to concerns about the COVID-19 pandemic. Overall, Exacerbation subjects (N=26) were 65 years old, predominantly male, former smokers, had a FEV_1_ of 26% predicted on inhaled bronchodilator therapy (Table 1). The majority of the Exacerbation subjects were GOLD stage 4 and all were GOLD risk group E. Exacerbation subjects reported having an average of 5.7 exacerbations in the past year, including 3.3 hospitalizations with respiratory failure and 2.4 that were treated as an outpatient. There were no differences between the 8 Exacerbation subjects who completed follow-up and the 18 subjects who did not (Table 1). The 8 Exacerbation subjects that completed follow up were similar to themselves at hospitalization (N=8), except for a lower heart rate (Table 1). Eight Stable COPD controls were enrolled and matched to Exacerbation patients for age, smoking status, and FEV_1_ % predicted. Baseline characteristics for Stable COPD controls (N=8) were similar to Exacerbation subjects at follow up (N=8), with the exception of prior year exacerbations, resting heart rate and GOLD risk group (Table 1).

**Table 1.** Subject Characteristics*.

| Characteristic | Exacerbation<br>During<br>Hospitalization<br>(A) | Exacerbation<br>During<br>Hospitalization<br>No Follow-Up<br>(B) | Exacerbation<br>During<br>Hospitalization<br>With Follow-Up<br>(C) | p-Value<sup>†</sup><br>B vs. C | Exacerbation<br>During<br>Follow-Up<br>(D) | Stable<br>COPD<br>Matched<br>(E) | p-Value<sup>†</sup><br>D vs. E |
| --- | --- | --- | --- | --- | --- | --- | --- |
| N | 26 | 18 | 8 |  | 8 | 8 |  |
| Age, y | 64.6 (8.2) | 65.5 (7.5) | 62.6 (10.0) | 0.40 | 62.6 (10.0) | 65.0 (9.6) | 0.60 |
| Female, n (%) | 8 (30.8) | 6 (33.3) | 2 (25.0) | >0.99 | 2 (25.0) | 3 (37.5) | >0.99 |
| **Race/Ethnicity** |  |  |  |  |  |  |  |
| White, n (%) | 20 (76.9) | 14 (77.8) | 6 (75.0) | >0.99 | 6 (75.0) | 8 (100) | 0.47 |
| Black, n (%) | 4 (15.4) | 4 (15.4) | 4 (15.4) |  | 1 (12.5) | 0 (0) |  |
| Other, n (%) | 1 (3.8) | 1 (3.8) | 1 (3.8) |  | 0 (0) | 0 (0) |  |
| Hispanic, n (%) | 1 (3.8) | 1 (3.8) | 1 (3.8) |  | 1 (12.5) | 0 (0) |  |
| **Smoking** |  |  |  |  |  |  |  |
| Current Smoker, n (%) | 11 (42.3) | 6 (33.3) | 5 (62.5) | 0.22 | 5 (62.5) | 2 (25.0) | 0.31 |
| Pack-years | 61.2 (45.7) | 62.3 (47.7) | 58.8 (43.8) | 0.80 | 71.8 (35.5) | 62.1 (26.8) | 0.75 |
| **Respiratory Medications** |  |  |  |  |  |  |  |
| SABA, n (%) | 23 (88.5) | 16 (88.9) | 7 (87.5) | >0.99 | 8 (100) | 7 (87.5) | >0.99 |
| LABA, n (%) | 22 (84.6) | 16 (88.9) | 6 (75.0) | 0.56 | 6 (75.0) | 6 (75.0) | >0.99 |
| LAMA, n (%) | 17 (65.4) | 10 (55.6) | 7 (87.5) | 0.19 | 7 (87.5) | 8 (100) | >0.99 |
| ICS, n (%) | 22 (84.6) | 16 (88.9) | 6 (75.0) | 0.56 | 6 (75.0) | 7(87.5) | >0.99 |
| Macrolide Antibiotic, n (%) | 6 (23.1) | 3 (16.7) | 3 (37.5) | 0.33 | 0 (0) | 0 (0) | >0.99 |
| PDE-4 Inhibitor, n (%) | 2 (7.7) | 2 (11.1) | 0 (0) | >0.99 | 0 (0) | 0 (0) | >0.99 |
| **Comorbidities** |  |  |  |  |  |  |  |
| Hypertension, n (%) | 13 (50.0) | 10 (55.6) | 3 (37.5) | 0.67 | 3 (37.5) | 2 (25.0) | >0.99 |
| Heart Failure, n (%) | 8 (30.8) | 6 (33.3) | 2 (25.0) | >0.99 | 2 (25.0) | 0 (0) | 0.47 |
| Atrial Fibrillation, n (%) | 3 (11.5) | 3 (16.7) | 0 (0) | 0.53 | 0 (0) | 0 (0) | >0.99 |
| **Past Year Exacerbations** |  |  |  |  |  |  |  |
| Hospital + RF | 3.3 (2.3) | 3.5 (2.6) | 2.9 (1.4) | 0.80 | 3.0 (1.5)** | 0.0 (0.0) | <0.001 |
| Outpatient | 2.4 (2.6) | 2.4 (3.0) | 2.4 (1.4) | 0.43 | 3.0 (1.9)** | 0.5 (0.8) | 0.01 |
| Total | 5.7 (4.1) | 5.9 (4.7) | 5.3 (2.2) | 0.73 | 6.0 (2.8)** | 0.5 (0.8) | <0.001 |
| **Vital Signs** |  |  |  |  |  |  |  |
| Heart Rate, per min | 107.2 (20.3) | 109.2 (21.1) | 102.9 (18.9) | 0.32 | 85.7 (8.3)<sup>†</sup> | 70.4 (11.4) | 0.02 |
| Respiratory Rate, per min | 23.9 (6.4) | 24.7 (6.5) | 22.1 (6.4) | 0.29 | 16.3 (2.4) | 15.5 (2.8) | 0.58 |
| SpO<sub>2</sub>, % | 93.1 (4.8) | 93.0 (5.0) | 93.4 (4.6) | 0.87 | 93.6 (3.3) | 95.5 (2.4) | 0.29 |
| BMI, units | 23.9 (6.5) | 24.4 (6.5) | 22.9 (6.7) | 0.52 | 24.2 (6.1) | 25.3 (5.5) | 0.69 |
| **Venous Blood Gas<sup>\*\*\*</sup>** |  |  |  |  |  |  |  |
| pH, units | 7.3 (0.1) | 7.3 (0.1) | 7.3 (0.1) | >0.99 |  |  |  |
| pCO<sub>2</sub>, mmHg | 63.1 (21.9) | 74.6 (21.1) | 50.7 (14.7) | 0.12 |  |  |  |
| **Lung Function** |  |  |  |  |  |  |  |
| FEV<sub>1</sub>, % predicted | 25.8 (8.6) | 24.9 (7.8) | 27.9 (10.4) | 0.33 | 31.7 (12.9) | 33.3 (16.0) | 0.82 |
| FEV<sub>1</sub>/FVC, ratio | 0.4 (0.1) | 0.4 (0.1) | 0.4 (0.1) | 0.30 | 0.4 (0.1) | 0.4 (0.1) | 0.42 |
| DLCO, % predicted | 36.1 (17.9) | 31.4 (15.6) | 46.5 (19.2) | 0.08 | 42.0 (18.0) | 41.3 (20.6) | >0.99 |
| **GOLD Stage** |  |  |  | 0.33 |  |  | 0.69<sup>‡</sup> |
| 2, n (%) | 0 (0) | 0 (0) | 0 (0) |  | 1 (14.3) | 1 (12.5) |  |
| 3, n (%) | 6 (23.1) | 3 (16.7) | 3 (37.5) |  | 4 (57.1) | 3 (37.5) |  |
| 4, n (%) | 20 (76.9) | 15 (83.3) | 5 (62.5) |  | 2 (28.6) | 4 (50.0) |  |
| **GOLD Risk Group** |  |  |  |  |  |  | 0.001 |
| B, n (%) | 0 (0) | 0 (0) | 0 (0) |  | 0 (0) | 7 (87.5) |  |
| E, n (%) | 26 (100) | 18 (100) | 8 (100) |  | 8 (100) | 1 (12.5) |  |

\*Data are presented as mean (SD) unless otherwise stated.
*Definition of abbreviations:* FEV<sub>1</sub> = forced expiratory volume in 1 second, FVC = forced vital capacity, DLCO = diffusion capacity of the lungs for carbon monoxide, VBG = venous blood gas, GOLD = Global Initiative for Chronic Obstructive Lung Disease
<sup>†</sup>Wilcoxon tests were used for continuous variables and Fisher's exact tests (or chi-square tests<sup>‡</sup>) were used for categorical variables.
<sup>\*\*</sup>Any additional exacerbations were added to past year exacerbations assessed in hospital.
<sup>‡</sup>p < 0.05 compared to the same subjects during hospitalization (column C vs D), using paired t-tests.
<sup>\*\*\*</sup>N=8 for those not followed-up, and 3 for those followed-up

**Figure 1.**
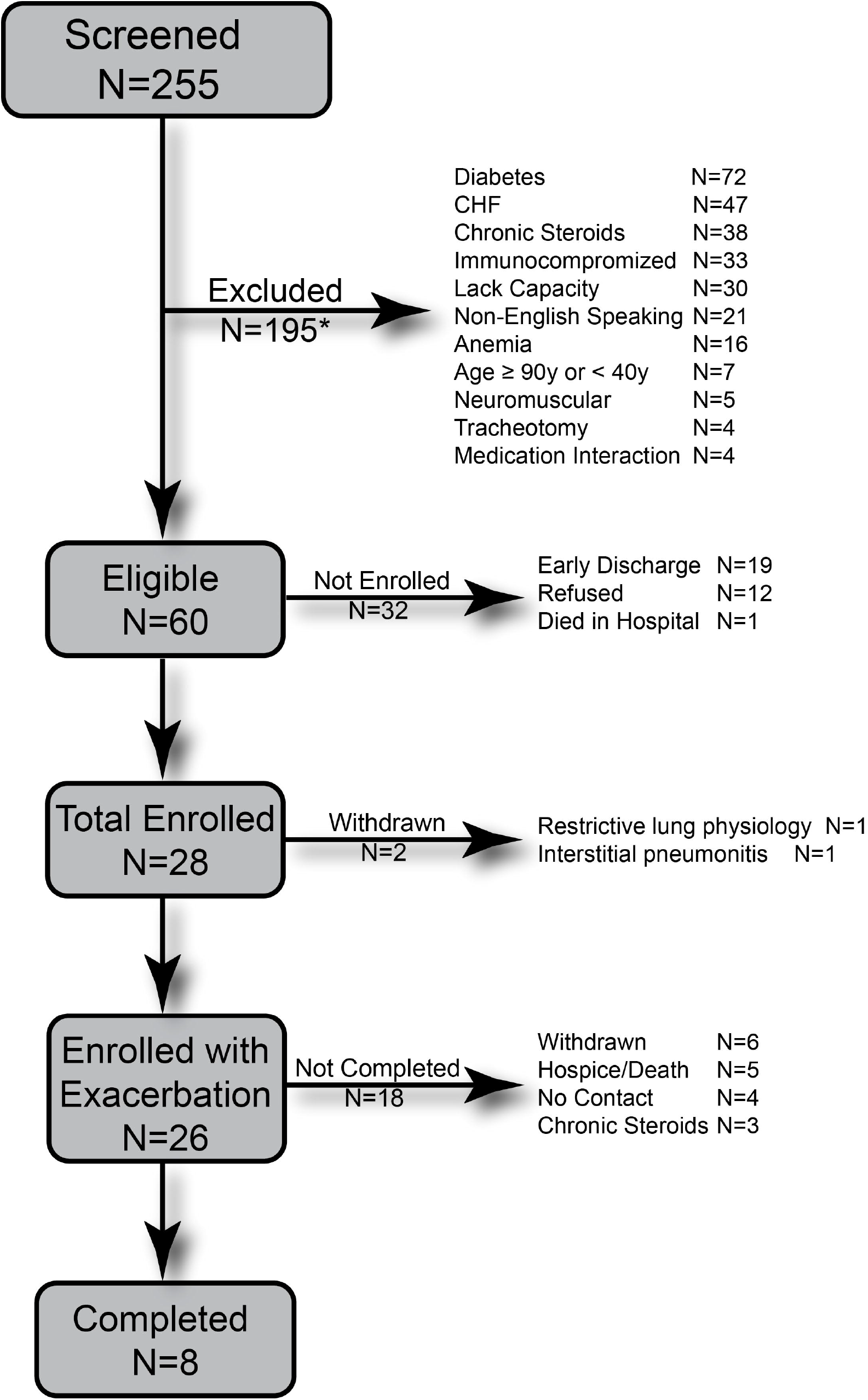
Patient Selection Flow Diagram. Study flow diagram showing screened, eligible, enrolled and completed subjects *Total exclusions (N=277) does not equal the total number of subjects excluded (N=195) due to subjects with multiple exclusions. Two enrolled subjects were removed after being found to have restrictive lung physiology and interstitial pneumonitis.

### *Ex Vivo* Corticosteroid Sensitivity

Corticosteroid sensitivity of the Exacerbation and Stable COPD groups was determined by examining differences in the methylprednisolone response curves and by calculating the IC_50_ of methylprednisolone to suppress IL-8 (primary outcome), IL-6, TNFα, and IL-1β in mononuclear cells and heparinized whole blood that had been stimulated with LPS *ex vivo*. Studies done in mononuclear cells and whole blood were consistent, in that they showed no difference in corticosteroid sensitivity across study groups as measured by differences in the methylprednisolone IC_50_ to suppress IL-8, IL-6, TNFα, and IL-1β or differences in the response curves or (Fig. 2).

**Figure 2.**
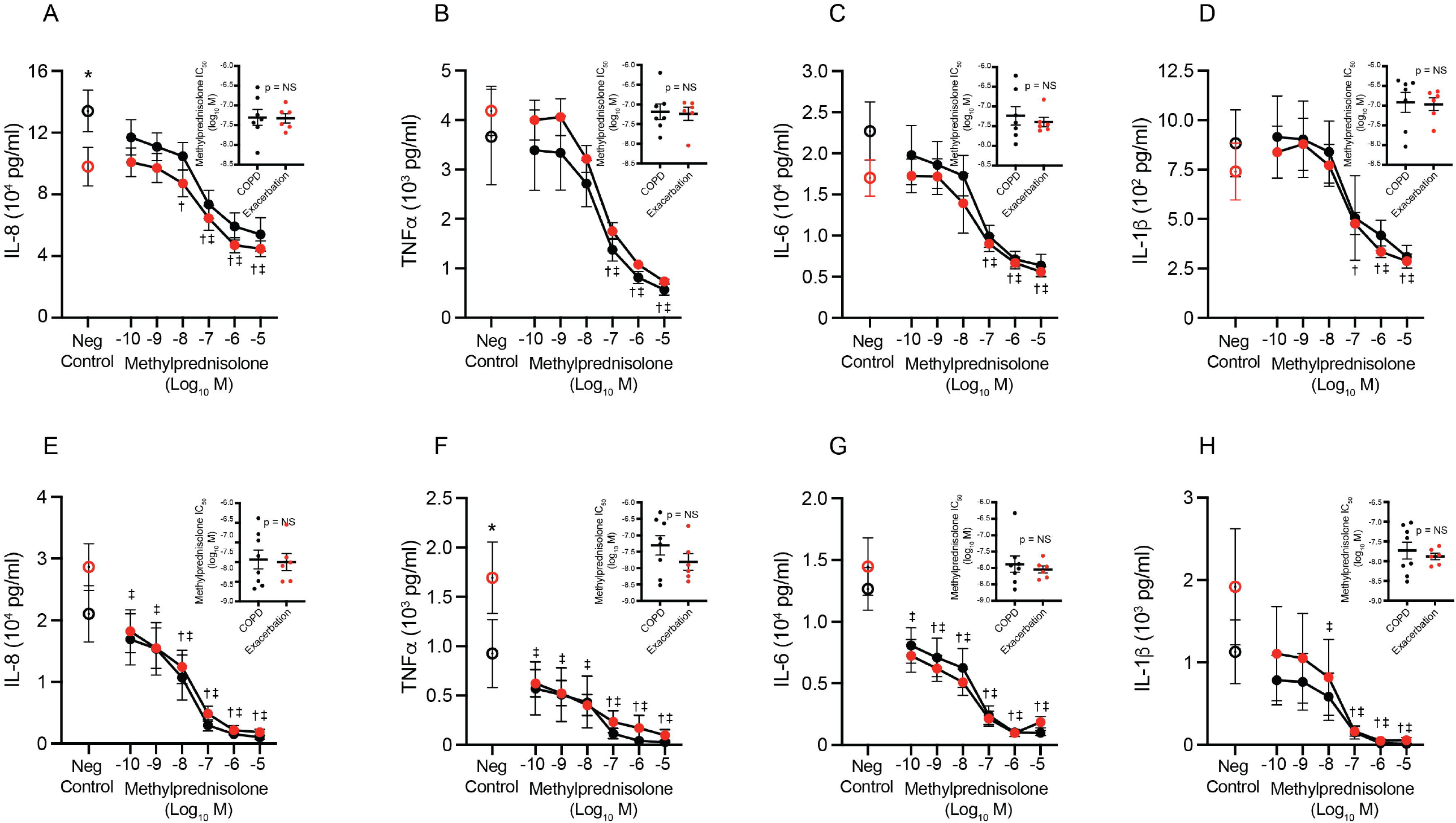
*Ex Vivo* Effects of Corticosteroids on Inflammatory Mediators. Panels A-H show the effects of methylprednisolone (10^−10^-10^−5^ M) on secretion of IL-8, TNFa, IL-6 and IL-1β by LPS-stimulated peripheral blood monocytes (A-D) and whole blood (E-H) from subjects following an acute exacerbation of COPD with respiratory failure (red circles) *versus* Stable COPD (black circles). Negative control was stimulated with LPS but was not treated with methylprednisolone. Figure inserts in panels show a comparison of methylprednisolone IC_50_ values between Exacerbation (red circles) and Stable COPD groups (black circles). p = NS for differences in methylprednisolone IC_50_ in panel inserts. ^†^p < 0.05 for indicated methylprednisolone concentration compared to Stable COPD negative control. ^‡^p < 0.05 for indicated methylprednisolone concentration compared to Exacerbation negative control. *p = 0.01 for a difference in IL-8 and TNFa concentration in the negative control for Exacerbation *versus* Stable COPD. For other panels, all comparisons between Exacerbation and Stable COPD for levels and changes at each concentration of methylprednisolone are nonsignificant.

### *In Vivo* Corticoid Sensitivity on Cells and Inflammatory Mediators

Corticosteroid responsiveness was determined *in vivo* by the ability of a single, 60 mg intravenous dose of methylprednisolone to suppress mononuclear cell numbers (Fig. 3) and to regulate pro- or anti-inflammatory mediators in the blood (Fig. 3) over 6 hours in Exacerbation *versus* Stable COPD cohorts. These studies showed that methylprednisolone decreased total mononuclear cells and monocytes, as has been demonstrated in prior studies[15, 19–21], and that there were no differences between Exacerbation and Stable COPD groups. Methylprednisolone also decreased lymphocytes in the Exacerbation and Stable COPD groups, with the notable difference that lymphocytes were higher in the Exacerbation group at baseline and declined more quickly in response to methylprednisolone compared to the Stable COPD group. In contrast, there were no differences in the ability of methylprednisolone to regulate pro- or anti-inflammatory mediators *in vivo* (Fig. 3).

**Figure 3:**
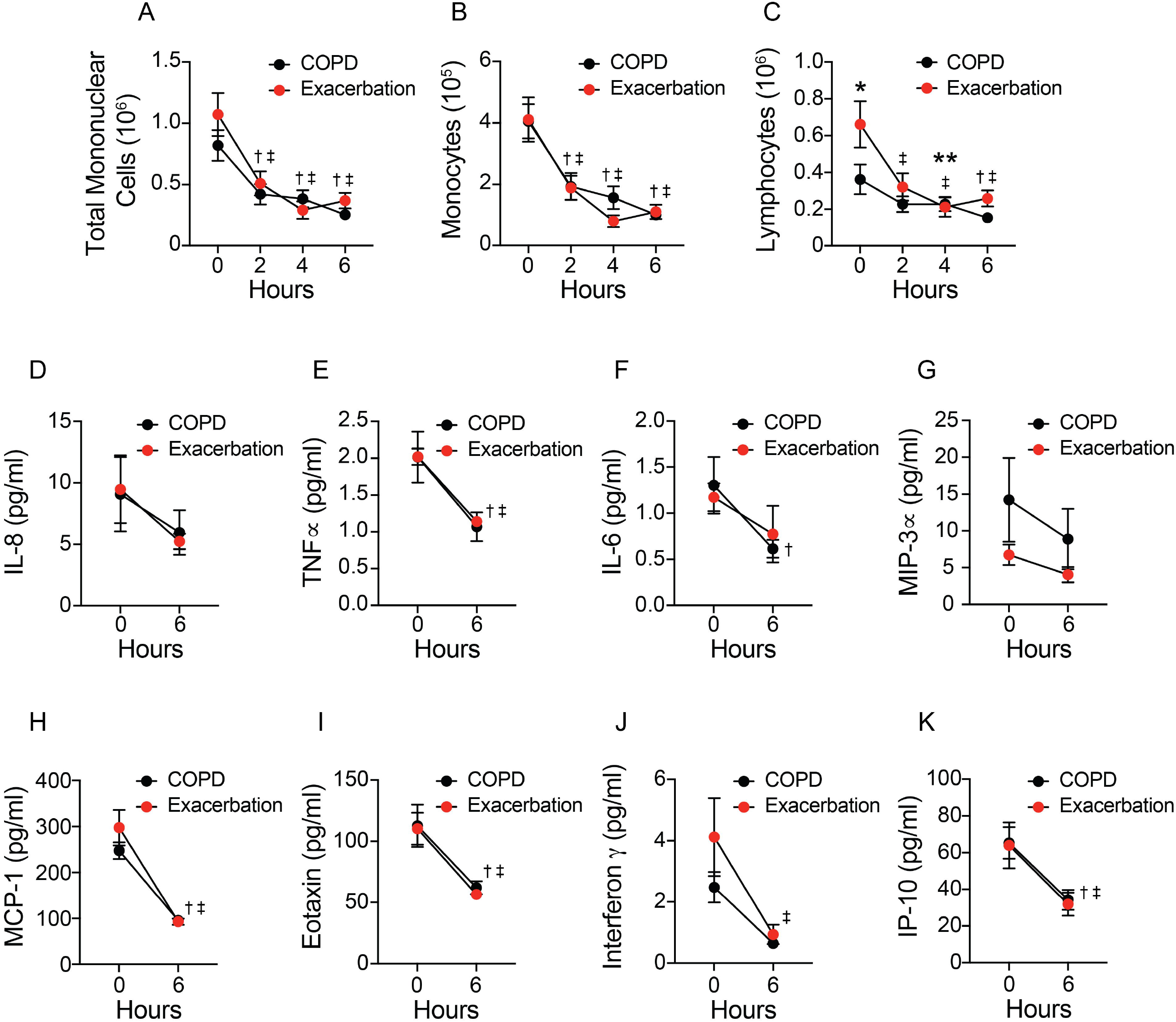
*In Vivo* Effects of Corticosteroids on Cell Counts and Inflammatory Mediators. Figure panels show the effects of 60 mg intravenous methylprednisolone on A) total peripheral blood mononuclear cells, B) monocytes, C) lymphocytes, D) IL-8, E) TNFa, F) IL-6, G) MIP-3a, H) MCP-1, I) Eotaxin, J) Interferon g and K) IP-10 over 6 hours. ^†^p < 0.05 for indicated time-point compared to Stable COPD at time = 0. ^‡^p < 0.05 for indicated time-point compared to Exacerbation at time = 0. *p = 0.002 for a difference in lymphocyte concentration at time = 0 for Exacerbation *versus* Stable COPD. **p = 0.02 for difference in change of lymphocyte concentration between 0 and 4 hours for Exacerbation *versus* Stable COPD. For other panels, all comparisons between Exacerbation and Stable COPD for levels and changes at each time point are nonsignificant.

### *In Vivo* Corticoid Sensitivity on Gene Expression

Finally, the study examined whether intravenous methylprednisolone had a differential effect on gene expression by mononuclear cells from Exacerbation *versus* Stable COPD subjects. A principal components analysis of gene expression by group, subject and time (0, 2, 4 and 6 hours) showed wide heterogeneity (Fig. 4A). RNA transcripts were normalized to baseline (0 hour) to correct for batch effects and differences in gene expression. Log2 of 2h/0h, 4h/0h and 6h/0h were used as input data. Data were fitted with repeated ANOVA (Ime4 library in R) for group + time + group x time analyses. Multiple-correction, FDR < 0.05 was applied to extract significant genes (N=396). When samples were normalized by baseline gene expression (0-2, 0-4 and 0-6 hours), principal component analysis showed clustering by individual subjects, but not by group or time (Fig. 4B). Group, time and group x time analyses of normalized data showed that methylprednisolone affected 6 genes by group analysis (Exacerbation *vs*. Stable COPD) with an FDR < 0.05, 396 genes by time analysis (*i*.*e*. 0-2, 0-4 and 0-6 hours), and 6 genes by group x time analyses (Fig. 4C; Supplementary Table 1). The group analysis identified differences in genes involved with protein folding (PPIAL4G), RNA decay (UPF1), RNA metabolism and gene regulation (RTCA-AS1), and 3 non-coding RNAs without known function. The 396 genes identified in the time analysis segregated into three clusters representing different patterns of expression over 6 hours, including 215 early response genes that increased at 2 hours, 26 middle response genes that increased at 4 hours, and 155 late response genes that increased at 6 hours (Fig. 4B and C). Methylprednisolone decreased genes associated with monocytes, T cells and IL-2 signaling, and increased genes associated with B lymphoblasts, NK cells, the cell cycle, G2-M checkpoint and mTORC1 signaling pathways. The group x time analysis identified five significant genes and one pseudogene (AKR7A2P1) that were differentially regulated in Exacerbation *versus* Stable COPD subject over time. These genes included POLR2J, a regulator of transcription that increased at 4 hours, AARS, a regulator of translation that increased at 4 hours and was also present in the middle response cluster of the time analysis, HYPK, a gene involved with protein stability that decreased at 4 hours, and two genes involved with protein degradation that increased (UBQLN4) of decreased at 4 hours (AFG3L2).

**Figure 4:**
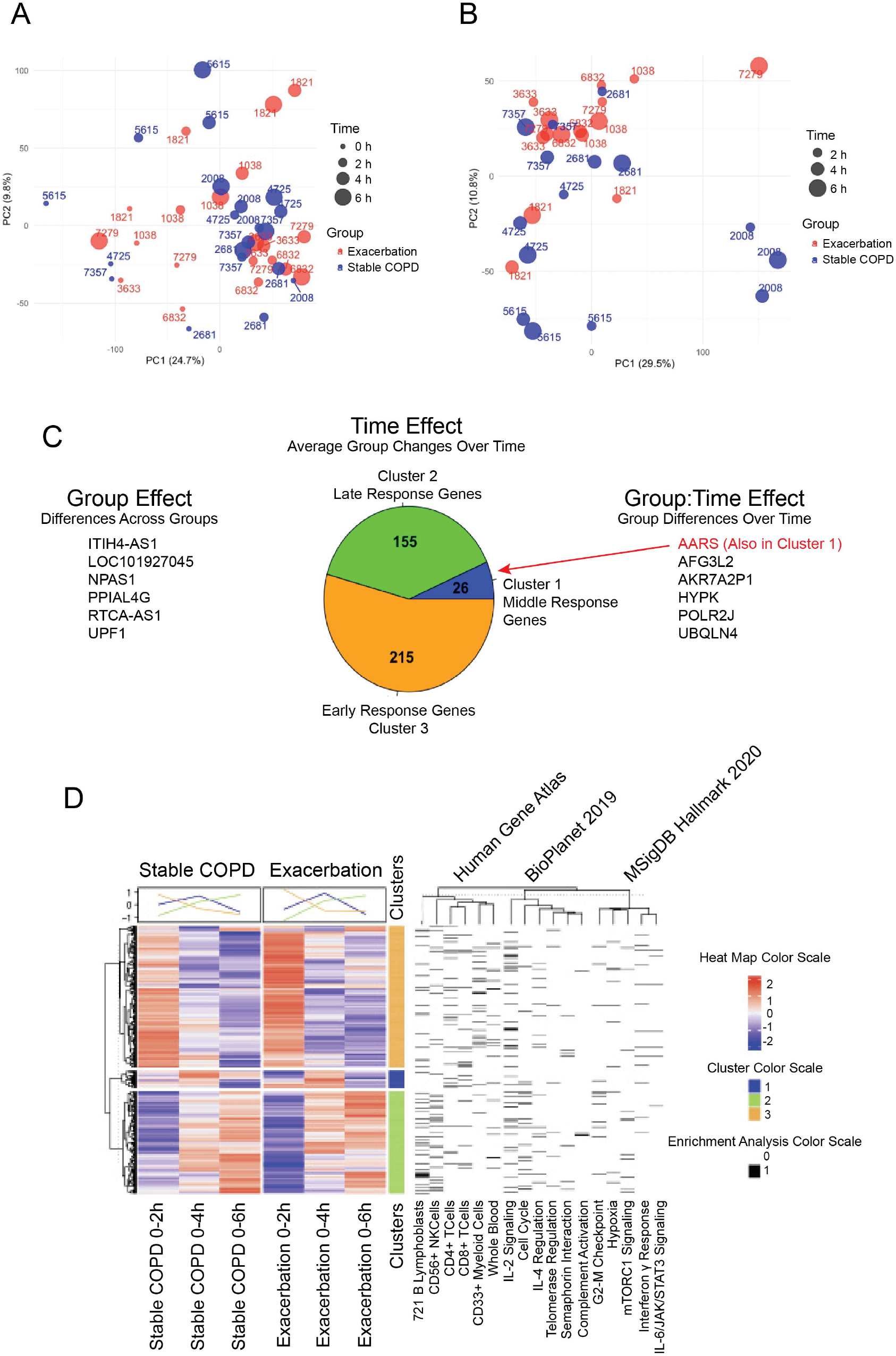
In Vivo Effects of Methylprednisolone on Gene Expression by Exacerbation and Stable COPD Normalized to Baseline. This figure shows the effects of 60 mg intravenous methylprednisolone on differential peripheral blood mononuclear cell (PBMC) gene expression at 0, 2, 4 and 6 hours for Exacerbation and Stable COPD subjects (N=5, each). Panels A and B show Principal Components Analysis (PCA) Plots for PBMC gene expression of Exacerbation (red) and Stable COPD (black) subjects (N=5, each) before (A) and after (B) normalization by gene expression by 0h. C) Effect of methylprednisolone on change of gene expression compared to baseline (0h) with an FDR < 0.05. Separate analyses were done to determine effects by Group (Exacerbation vs. Stable COPD), Time (0-2h, 0-4h and 0-6h) and Group x Time. Group analysis identified 6 differentially expressed genes. Time analysis identified 396 differentially expressed genes that clustered as early (orange; 2h), middle (blue; 4h) and late (green; 6h) response genes *versus* time 0h. The Group x Time analysis identified 6 differentially expressed genes, including AARS (shown in red), which was also present in Cluster 1 of the Time analysis (see red arrow). D) The Time analysis heatmap shows the average change of gene expression for 0-2h, 0-4h and 0-6h for Exacerbation and Stable COPD subjects, which clustered by expression pattern (top of heat map). The Human Gene Atlas, BioPlanet 2019 and mSigDB Hallmark 2020 were used for cell type and pathways analysis of each cluster.

## Discussion

Clinical studies have consistently shown that lower doses and shorter courses of corticosteroids are sufficient to treat people with exacerbations in both outpatient and inpatient settings [13–15]. In contrast, people who are admitted to the intensive care unit with an exacerbation continue to be treated with higher doses of corticosteroids that range from 3 – 12 times higher than is recommended for people hospitalized without respiratory failure, despite evidence that higher doses are associated with more side effects[7, 22]. This difference in corticosteroid dosing may be due to a paucity of studies that have evaluated dosing in exacerbations with respiratory failure[17, 18, 23, 24] and/or the impression that people who have respiratory failure have intrinsic corticosteroid resistance that requires higher dosing[22]. This study was designed to test the hypothesis that people who suffer from exacerbations with respiratory failure have intrinsic steroid resistance that is part of their endotype at baseline during a period of quiescence. In contrast, the study found no evidence of corticosteroid resistance to suppress inflammation in people following an exacerbation with respiratory failure, suggesting that lower doses of corticosteroids may be as effective as higher doses that are frequently used today.

This study used both *ex vivo* and *in vivo* methods to test the study hypothesis. These studies found that methylprednisolone suppressed LPS-induced cytokines in mononuclear cells and heparinized whole blood to an equal degree in Exacerbation and Stable COPD groups. It was important to perform these experiments in both heparinized whole blood and in mononuclear cells, because they contain different leukocyte components. Whole blood contains a mixed population of granulocytes (*e*.*g*. neutrophils, eosinophils and basophils), lymphocytes and monocytes, while mononuclear cells contain only lymphocytes and monocytes. Neutrophils make up 55 - 70% of leukocytes in heparinized whole blood, and they are known to have a variable response to corticosteroids[25]. On the other hand, corticosteroids rapidly cause eosinophil apoptosis [26] and studies suggest that corticosteroids may be more effective for treatment of exacerbations in people with higher blood eosinophils[27, 28]. The presence of eosinophils could have been the reason that 10^−10^ M methylprednisolone suppressed LPS-induced cytokine production in heparinized whole blood compared to mononuclear cells. Using data from the medical record, Exacerbation and Stable COPD subjects had equal numbers of blood eosinophils (411 ± 254 cells per microL in Exacerbation subjects vs. 462 ± 472 cells per microL in Stable controls; p = 0.79). Despite these differences in cell components, neither mononuclear cells nor heparinized whole blood demonstrated any differences in sensitivity to methylprednisolone for the Exacerbation *versus* Stable COPD groups.

This study found that 60 mg of intravenous methylprednisolone decreased mononuclear cells, plasma inflammatory mediators, and altered mononuclear cells gene expression to the same degree in Exacerbation and Stable COPD groups. These results are consistent with other studies that demonstrated the effects of intravenous corticosteroids on plasma inflammatory mediators, mononuclear cell counts and gene expression in normal subjects [19–21]. In contrast, blood lymphocytes were present in higher numbers and decreased more steeply in the Exacerbation group compared to Stable COPD controls. The presence of lymphocytosis in Exacerbation subjects suggests that they have chronic immune activation ~3 months post hospitalization. We did not assess potential causes of persistent lymphocytosis, or the type/activation status of blood lymphocytes. The study also showed that genes involved with transcription (POLR2J), translation (AARS) and protein stability (HYPK) and protein degradation (UBQLN4 and AFG3L2) were differentially expressed in the group x time analysis. While few-in-number, these genes are involved with fundamental cellular processes that could have important consequences for the exacerbation phenotype. Taken together, results indicate that immune dysregulation and select differences in corticosteroid responsiveness are present in people who have had exacerbations with respiratory failure, but there is no evidence for steroid resistance.

The study has several limitations including the small number of subjects that completed the study.

This was primarily due to exclusion criteria needed to avoid adverse effects from intravenous methylprednisolone (Fig. 1). Ongoing severe illness, recurrent hospitalization, prednisone use, hospice and death also made it extraordinarily difficult for Exacerbation subjects to complete the study. Despite these barriers, data clearly showed no evidence for steroid resistance. Another limitation is that corticosteroid sensitivity was not tested in airway epithelial cells. The Exacerbation group had severe airflow limitation, which precluded safe research bronchoscopy. Nasal epithelial cells could have been tested instead, but it is not certain that their response to corticosteroids genuinely reflects lower airway biology. It is noteworthy that similar methods have been used to demonstrate the presence of systemic corticosteroid resistance in asthma patients who are clinically resistant to corticosteroids[29], implying that results of systemic corticosteroid sensitivity may mirror biology present in the airways. Finally, the study did not look for steroid resistance during an active exacerbation with respiratory failure, because it would not examine the underlying endotype and would not be practical.

Exacerbations with respiratory failure are a devastating complication of COPD[5, 8, 11]. Despite the fact that corticosteroids have been a cornerstone of exacerbation therapy for over 40 years[30], we still do not know the best dose or length of treatment in those who are admitted to the intensive care unit with respiratory failure[7, 17, 18, 23]. We do know from epidemiology studies, however, that high doses of corticosteroids are associated with decreased efficacy and increased harm to this group[7]. This knowledge gap has occurred because randomized controlled studies of steroids for this vulnerable group have not been performed outside of a placebo-controlled design[17, 18, 24]. As a result, there is a wide diversity of opinion about the optimal corticosteroid dose for exacerbations with respiratory failure[22], with some studies even questioning whether corticosteroids have any benefit at all[17, 24, 31]. As James H. Shelhamer, MD once said to critical care medicine fellows at NIH, “strong differences of opinion in medicine are usually due to a lack of knowledge”. Given the uncertainty that we face today regarding corticosteroid dosing for exacerbations with respiratory failure, it is critical that studies are performed to answer this question.

The most important message from this study is that there is no evidence that the endotype of COPD exacerbations with respiratory failure is defined by corticosteroid resistance to suppress inflammation, suggesting that high doses of corticosteroids may not have greater benefit than lower doses, and might even cause harm[7, 22]. Corticosteroids do have differential effects on a small group of genes that affect fundamental cellular processes, but, as yet, the significance of these findings for exacerbations with respiratory failure is unknown. Perhaps as important, the recognition that lymphocytosis persists three months after an exacerbation, suggests that chronic immune activation may play an important role in recurrent episodes of respiratory failure and should be a target for further research.

## Data Availability

All data produced in the present study are available upon reasonable request to the authors

## Author Acknowledgements

The authors would like to thank Dr. James H. Shelhamer for a lifetime of superb teaching, for his astute observations of clinical medicine, and for his critical reading of the manuscript.

## Take Home Message

People who suffer from COPD exacerbations with respiratory failure have persistent lymphocytosis, an altered gene response to corticosteroids, but do not have intrinsic corticosteroid resistance to suppress inflammation.

