## Supplementary material for "Corticosteroid Responsiveness Following COPD Exacerbations with Respiratory Failure": Online Data Supplement

**Methods**

**Patient Population.** Exacerbation subjects were recruited during their index hospitalization and were invited to return for an outpatient visit for testing ≥45 days after hospital discharge. Matched, Stable COPD controls were recruited from the COPD and Lung Transplantation Clinics. Inclusion criteria for the exacerbation cohort were 1) emergency department or intensive care unit physician diagnosis of a COPD exacerbation, 2) age ≥ 40 years, 3) need for non-invasive or invasive ventilator support in the emergency department or intensive care unit during the first 24 hours of hospitalization. Inclusion criteria for the Stable COPD cohort were 1) physician diagnosis of COPD and 2) age ≥ 40 years. Stable COPD patients were also frequency-matched to exacerbation patients for age (± 10 year increments), current/former smoking status (former smoker = no smoking for ≥ 1 month) and lung function (FEV_1_% predicted by ± 10% increments). The study excluded people in the Stable COPD group if they had a moderate exacerbation treated as an outpatient or through the emergency department within 6 months, or a severe exacerbation treated as an inpatient within 12 months. Subjects in both cohorts were excluded for 1) systemic steroid or antibiotic use ≤ 30 days prior to the return visit, 2) hemoglobin < 8.0 g/dl, 3) acute pulmonary embolism, 4) history of interstitial lung disease or heart failure with respiratory exacerbation, 5) age ≥ 90 years, 6) known pregnancy, 7) nursing mothers and 8) incarcerated individuals. People were also excluded for a history of diabetes, immunodeficiency, tracheostomy, or neuromuscular disease because of increased risk of hyperglycemia, immunosuppression and muscle weakness related to intravenous methylprednisolone. Likewise, people were excluded if they were taking drugs that induce cytochrome P450 3A enzyme activity and affect corticosteroid metabolism.

**Study Design***.* We conducted a matched, case-control study to examine steroid sensitivity following COPD exacerbations with respiratory failure from October 2017 through June 2020 (Clinical Trials.gov ID: NCT03680495. The Colorado Multiple Institutional Review Board approved the study (COMIRB 16-0256) and all subjects completed written, informed consent.

**Measurements.**

To examine the hypothesis that systemic steroid resistance is present in patients with a history of exacerbations with respiratory failure, we initiated a pilot study of Exacerbation patients *versus* Stable COPD controls with *ex vivo* and *in vivo* outcomes. The primary outcome was steroid resistance measured by a difference in the *ex vivo* IC_50_ of methylprednisolone to suppress release of IL-8 by peripheral blood mononuclear cells (mononuclear cells) stimulated with LPS in Exacerbation *versus* Stable COPD groups. The *ex vivo* IC_50_ of methylprednisolone to suppress release of mediators was tested in mononuclear cells and heparinized whole blood for IL-8, IL-6, TNFα and IL-1β. *In vivo* studies were also performed to test for the presence of steroid resistance. These studies examined the ability of a single, intravenous dose of methylprednisolone to change plasma mediators, PBMC counts and gene expression over 6 hours in Exacerbation *versus* Stable COPD groups.

*Ex Vivo Studies of Steroid Resistance.* Mononuclear cells were collected using citrated CPT tubes (BD Biosciences, San Jose, CA 95131). Total and differential cell counts were performed on mononuclear cells to assess lymphocyte and monocyte components and to confirm that lack of neutrophil contamination. Mononuclear cells were resuspended in X-vivo 10 media (Lonza Bioscience; Allendale, NJ) plus 10% pooled, heat inactivated human serum (Atlanta Biologicals, Lawrenceville, GA, 30043, ) at 1x10^6^ cells/ml, aliquoted at 100µl per well of a 96-well plate, treated with methylprednisolone at 0, 10^10^, 10^9^, 10^8^, 10^7^, 10^6^, and 10^5^ M (Sigma-Aldrich, Saint Louis, MO, 63103) for 1 hour and stimulated with *E. coli* O11B4 LPS (List Labs, Campbell, CA, 95008) at 10 ng/ml for 24h. Supernatants were collected, assessed for concentrations of IL-8, IL-6, TNFα and IL-1β (Meso Scale Diagnostics, Rockville, MD 20850) to determine the methylprednisolone IC_50_.

Whole blood was collected in heparinized tubes (Sarstedt, Numbrecht, Germany), aliquoted in 48 well plates at 300 µl per well, treated with methylprednisolone at 0, 10^10^, 10^9^, 10^8^, 10^7^, 10^6^ and 10^5^ M (Sigma-Aldrich, Saint Louis, MO, 63103) for 1 hour and stimulated with *E. coli* O11B4 LPS (List Labs, Campbell, CA, 95008) at 10 ng/ml for 24h. Supernatants were collected, assessed for concentrations of IL-8, IL-6, TNFα and IL-1β (Meso Scale Diagnostics, Rockville, MD) to determine the methylprednisolone IC_50_.

*In Vivo Studies of Steroid Resistance.* *In vivo* studies of steroid resistance were performed by testing the ability of methylprednisolone to suppress pro-inflammatory cytokines (IL-8, TNFα, IL-6, IFN-γ, IL-5) and chemokines (MIP-3α, MCP-1, MCP-4, IP-10, Eotaxin), and to activate the anti-inflammatory mediator, IL-10 (Meso Scale Diagnostics, Rockville, MD 20850) in serum at 0 and 6 hours post intravenous injection with 60 mg of methylprednisolone (Sigma-Aldrich, Saint Louis, MO, 63103). Group differences on total mononuclear cells, monocyte and lymphocyte cell counts were assessed in mononuclear cells collected at 0, 2, 4 and 6 hours following intravenous methylprednisolone.

*RNA Isolation and Sequencing.* Mononuclear cells were isolated using citrated CPT™ tubes (BD Biosciences, San Jose, CA 95131) and total RNA was extracted using Qiagen RNeasy Micro kit. Library prep and sequencing was done in the Genomics Shared Resource using Illumina NovaSeq 6000 sequencer for 2 × 150 cycles (paired-end reads). Data analysis involved removal of low quality reads, and trimming of the adapter sequence using Cutadapt ^1^. The raw sequencing reads were mapped against the Human reference genome hg38 using the HiSAT2/Cufflinks workflow as previously described.^2 3^ Transcripts were quantified using reads per kilobase per million mapped reads (RPKM), and RPKM from all samples were quartile-normalized before downstream analysis. The temporal pattern of transcriptional changes in the Exacerbation and Stable COPD groups were determined using a linear mixed-effects model, accounting for group, time, and their interaction, while adjusting for patient-specific variability.  Gene set enrichment analysis was performed using Human_Gene_Atlas, BioPlanet_2019, and MSigDB_Hallmark_2020 as the reference databases.^4^ Pathways with FDR <0.05 were considered significant.

*Clinical Measurements.* We collected personal and disease related information from all subjects, including demographics, the number and severity of exacerbations within the past year, outpatient respiratory medications, and hospital data for exacerbation patients. In the Exacerbation group, we also recorded the number and severity of exacerbations that occurred between the hospitalization and follow up. Post bronchodilator spirometry and diffusion capacity of the lungs for carbon monoxide (DLCO) was measured in the Exacerbation and Stable COPD groups at enrollment and during follow up using the ndd EasyOne Pro® LAB (ndd Medical Technologies, Andover, MA 01810).

*Data Analysis.* Statistical analyses were performed using SAS for Windows, version 9.4 (SAS Institute, Cary, NC). Comparisons between study subjects in the Exacerbation and Stable COPD groups were made (using t-tests or Wilcoxon tests if the distributions were skewed) for continuous variables and chi-square tests (or Fisher’s exact tests if cell sizes were small) for categorical variables. Comparisons involving longitudinal curves were carried out using linear models. All tests were 2-sided with a significance level of p < 0.05. All graphs show standard error.
